# Sex and Race Differences in the Performance of the European Society of Cardiology 0/1-Hour Algorithm with High-Sensitivity Troponin T

**DOI:** 10.1101/2023.04.28.23289294

**Authors:** Michael W. Supples, Anna C. Snavely, James C. O’Neill, Nicklaus P. Ashburn, Brandon R. Allen, Robert H. Christenson, Richard Nowak, R. Gentry Wilkerson, Bryn E. Mumma, Troy Madsen, Jason P. Stopyra, Simon A. Mahler

**Author notes:** Corresponding Author: Michael W. Supples, MD, MPH Department of Emergency Medicine Wake Forest School of Medicine Medical Center Boulevard, Winston-Salem, NC 27157 USA.

## Abstract

**Background:** The diagnostic performance of the high-sensitivity troponin T (hs-cTnT) European Society of Cardiology (ESC) 0/1-hour algorithm in sex and race subgroups is unclear, particularly in U.S. Emergency Department (ED) patients.

**Methods:** A pre-planned subgroup analysis of the STOP-CP cohort study was conducted. Participants with 0- and 1-hour hs-cTnT measures (Roche Diagnostics; Basel, Switzerland), prospectively enrolled at eight U.S. EDs from 1/2017-9/2018, were stratified into rule-out, observation, and rule-in zones using the hs-cTnT ESC 0/1 algorithm. The primary outcome was adjudicated 30-day cardiac death or MI. Rates of patient stratification to each ESC 0/1 zone and the proportion with 30-day cardiac death or MI in each zone were compared between subgroups with Fisher’s-exact tests. The negative predictive value (NPV) of the ESC 0/1 rule-out zone for 30-day cardiac death or MI was calculated and compared between subgroups using Fisher’s exact tests.

**Results:** Of the 1422 patients enrolled, 54.2% (770/1422) were male and 58.1% (826/1422) white with a mean age of 57.6 ± 12.8 years. At 30 days, cardiac death or MI occurred in 12.9% (183/1422) of participants. The ESC 0/1-h algorithm ruled-out more women than men [66.9% (436/652) vs 50.0% (385/770); p<0.001] and a similar proportion of white vs non-white patients [59.3% (490/826) vs 55.5% (331/596); p=0.16]. Among patients stratified to the rule-out zone, 30-day cardiac death or MI occurred in 1.1% (5/436) of women vs 2.1% (8/436) of men (p=0.40) and 1.2% (4/331) of non-white patients vs 1.8% (9/490) of white patients (p=0.58). The NPV for 30-day cardiac death or MI was similar among women vs men [98.9% (95%CI 97.3-99.6) vs 97.9% (95%CI 95.9-99.1); p = 0.40] and among white vs non-white patients [98.8% (95%CI 96.9-99.7) vs 98.2% (95%CI 96.5-99.2); p=0.39].

**Conclusions:** The ESC 0/1-h hs-cTnT algorithm ruled-out more women than men, but achieved similar NPV for 30-day cardiac death or MI in all subgroups. NPVs <99% in each subgroup suggests the hs-cTnT ESC 0/1-h algorithm may not be safe for U.S. ED use.

**Trial Registration:** High-Sensitivity Cardiac Troponin T to Optimize Chest Pain Risk Stratification (STOP-CP; ClinicalTrials.gov: NCT02984436; https://clinicaltrials.gov/ct2/show/NCT02984436)

**What is Known:**

- The European Society of Cardiology high-sensitivity troponin T (hs-cTnT) 0/1-hour algorithm has been validated as a tool to rule-out myocardial infarction (MI).
- Evaluations of its performance differences based on sex and race are limited.

**What the Study Adds:**

- In this multisite U.S. cohort study of 1422 adults, the high-sensitivity troponin T (hs-cTnT) ESC 0/1-hour algorithm had a similar NPV in men vs women and white vs non-white patients but did not achieve ≥99% NPV for 30-day cardiac death or MI in any subgroup.
- Emergency providers should be aware of the potential limitations of using the ESC 0/1-hour hs-cTnT algorithm for risk stratification because it may not satisfactorily exclude 30-day cardiac death or MI in any subgroup of ED patients (men, women, white patients, or non-white patients) with acute chest pain.

## INTRODUCTION

Chest pain leads to over 6.5 million Emergency Department (ED) visits in the U.S. per year.^1–3^ To optimize care for these patients, accelerated diagnostic protocols (ADP) have been developed to objectively guide patient risk stratification. The high sensitivity cardiac troponin T European Society of Cardiology 0/1-hour algorithm (hs-cTnT ESC 0/1-hour algorithm) is a commonly used ADP and is recommended by European and U.S. guidelines.^4–11^

In several studies, mostly conducted in Europe and Australasia, the hs-cTnT ESC 0/1-hour algorithm has demonstrated a ≥99% negative predictive value (NPV) for 30-day myocardial infarction (MI) or cardiac death.^6, 7, 12, 13^ Its performance in U.S. patients with acute chest pain is not as well established.^14^ In the primary analysis of the STOP-CP multisite U.S. cohort, the hs-cTnT ESC 0/1-hour algorithm did not achieve a 99% NPV, which is the threshold commonly used for ADP safety.^15^ However, it is unclear whether diagnostic performance was consistent across key patient subgroups, such as men, women, white patients, and non-white patients and there is a paucity of prior data evaluating sex or race differences in algorithm performance. Prior studies have demonstrated significant differences in 99^th^ percentile hs-cTnT values among men versus women and white patients versus non-white patients.^16–19^ In addition, it has been hypothesized that greater racial diversity of U.S. ED patients may explain differences in the diagnostic performance of the ESC 0/1-hour algorithm in the U.S. vs Europe and Australasia. Thus, it is plausible that performance of the ESC 0/1-hour algorithm may differ based on demographics.

The primary objective of this study was to evaluate and compare the diagnostic performance of the hs-cTnT ESC 0/1-hour algorithm for 30-day cardiac death or MI in men vs women and white vs non-white patients within the STOP-CP cohort. A secondary objective was to evaluate and compare diagnostic performance for 30-day major adverse cardiovascular events (MACE; defined as cardiac death, MI, or coronary revascularization) in these subgroups.

## METHODS

### Study Design and Setting

This is a preplanned subgroup analysis of the High-Sensitivity Cardiac Troponin T (Gen 5 STAT assay) to Optimize Chest Pain Risk Stratification (STOP-CP; ClinicalTrials.gov: NCT02984436) prospective, multicenter cohort study. STOP-CP enrolled patients with symptoms concerning for ACS at eight U.S. EDs from 01/25/2017 to 09/06/2018. Study sites included the University of Florida, Gainesville, Florida; Wake Forest University, Winston Salem, North Carolina; Henry Ford Health System, Detroit, Michigan; University of Maryland St. Joseph Medical Center, Towson, Maryland; University of Maryland Medical Center, Baltimore, Maryland; University of Maryland Baltimore Washington Medical Center, Glen Burnie, Maryland; University of California-Davis, Davis, California; and University of Utah, Salt Lake City, Utah. Institutional review board approval was obtained at all sites. Written informed consent was obtained for enrollment. STOP-CP methods are previously described.^20^ The Standards for Reporting of Diagnostic Accuracy Studies (STARD) guidelines helped direct the research and manuscript development processes.^21^

### Study Population

We prospectively enrolled ED patients ≥21 years of age with serial troponins ordered for the evaluation of possible acute coronary syndrome (ACS). Exclusion criteria included ST-elevation myocardial infarction, systolic blood pressure <90 mmHg, life expectancy <90 days, a non-cardiac illness requiring admission, inability to provide consent or be contacted for follow-up, non-English speaking, pregnancy, being a prisoner, or previous enrollment in the study.

### Data Collection

Serial blood samples were collected for hs-cTnT measurement at baseline (<1 hour from first clinical blood draw) and 1-hour later in lithium heparin tubes. hs-cTnT was quantified with the Gen 5 STAT assay on the Cobas e 601 analyzer (Roche Diagnostics; Basel, Switzerland). The assay has a range of 3 to 10,000 ng/L, limit of quantification at 6 ng/L, and a 99^th^ percentile upper reference limit (URL) of 19 ng/L in the U.S. with a coefficient of variation of <10%.^22^ Treating providers were blinded to hs-cTnT results therefore, patient care was dictated by local standards of care and guided by contemporary cTn results.

Demographic data were collected by research staff from the patient by self-report and were supplemented by the patient’s electronic medical record. These included age on the day of emergency department visit, sex, race and ethnicity, and risk factors (current or prior tobacco use, hypertension, hyperlipidemia, diabetes, family history of coronary artery disease (CAD), obesity, prior cerebrovascular accident, peripheral vascular disease, and end-stage renal disease). Initial electrocardiogram (ECG) findings of acute ischemia were indicated by the treating physician.

### ESC 0/1-Hour Algorithm

In each patient, hs-cTnT measures were used to stratify patients into rule-out, observation, and rule-in zones using established assay-specific cut-points.^4, 5, 20^ However, the hs-cTnT ESC 0/1-hour algorithm’s 0-hour rule-out cut-point of 5 ng/L (the limit of detection) was modified to 6 ng/L (the limit of quantification), because the U.S. Food and Drug Administration does not allow reporting below the limit of quantification. Based on prior derivation and validation studies, patients stratified to the rule-out zone were expected to have ≥99% NPV for cardiac death or MI.^6, 7, 13, 23^

### Outcomes

The primary outcome was 30-day cardiac death or MI, inclusive of index visit events. Secondary outcomes included: 1) 30-day MACE, 2) the individual MACE components (cardiac death, MI, and coronary revascularization) at index and from index through 30-days, and 3) efficacy, defined as the proportion of patients classified into the rule-out zone.^24, 25^ Medical record review and telephone follow-up through 30-days were completed to determine outcomes. Expert reviewers adjudicated any patient who experienced death, a clinical diagnosis of MI, or had an elevated contemporary cTn. Adjudicators classified deaths as cardiac or non-cardiac based on the Action to Control Cardiovascular Risk in Diabetes (ACCORD) trial definition, except for death due to stroke which was classified as a non-cardiac death.^26^ If the cause of death could not be determined, it was considered cardiac. MI was determined by the Fourth Universal Definition of MI: rise and fall of troponin (with at least one value ˃99^th^ percentile URL) with symptoms of ischemia, ECG evidence of ischemia, imaging evidence of new non-viable myocardium, a new regional wall motion abnormality, or evidence of thrombus on angiography.^27^

### Statistical Analysis

Counts, percentages, means and standard deviations, or medians and interquartile (IQR) ranges were used to describe the study population. To evaluate the performance of the ESC 0/1-hour algorithm, sensitivity, specificity, negative and positive predictive values (NPV and PPV), and negative and positive likelihood ratios (-LR and +LR) for 30-day cardiac death or MI and 30-day MACE were calculated. For efficacy, sensitivity, specificity, NPV and PPV, exact 95% confidence intervals (95%CI) were computed, and Fisher’s exact tests were used to compare by sex (men vs. women) and race (white vs. non-white patients). Likelihood ratios were calculated and reported with 95%CIs using the method of Simel et al.^28^ The asymptotic hypothesis test developed by Luts et al. was used to compare LRs by sex and race.^29^ Consistent with prior studies, sensitivity, NPV, and -LR were calculated for the rule-out zone (i.e., rule-out vs rule-in or observation) and specificity, PPV, and +LR were calculated for the rule-in zone (i.e., rule-in vs. observation or rule-out).^6, 7, 20, 30, 31^ Fisher’s exact tests were used to compare cardiac death or MI and MACE at index and from index though 30-days among sexes and races.

To assess the association of sex or race with index and 30-day cardiac death or MI and 30-day MACE, multivariable logistic regression was performed. Models were adjusted for age, sex, race (white vs. non-white patients), hypertension, diabetes, hyperlipidemia, obesity (BMI ≥ 30kg/m^2^), current smoking, prior stroke, peripheral vascular disease, and end stage renal disease. These variables were selected due to their relevance and inclusion in previous cardiovascular risk stratification work.^31^ Ethnicity was not included in this analysis. For each outcome we computed two separate adjusted models, one containing an interaction term between ESC 0/1-hour zone and sex, and another containing an interaction term between ESC 0/1-hour zone and race. Separate, unadjusted logistic regression models were fit to evaluate the association between sex and race with the primary and secondary outcomes within each ESC 0/1-hour zone. Unadjusted or adjusted odds ratios (aOR) with corresponding 95%CIs were calculated as appropriate for each logistic model.

## Results

This preplanned subgroup analysis included 1,422 patients. The patient flow diagram is shown in Figure 1. The cohort was 54.1% (770/1422) male and 58.1% (826/1422) white, with a mean age of 57.6±12.8 years. Patient demographics are presented in Table 1. The rate of 30-day cardiac death or MI in the cohort was 12.9% (183/1422), occurring in 8.7% (57/652) of women compared to 16.4% (126/770) of men (p < 0.001) and 11.2% (67/596) of non-white patients compared to 14.0% (116/826) of white patients (p = 0.12). MACE at 30-days was also more common in men than women [18.2% (140/770) vs. 9.7% (63/652); p < 0.001] and among white patients than non-white patients [16.0% (132/826) vs. 11.9% (71/596); p = 0.03].

**Figure 1.**
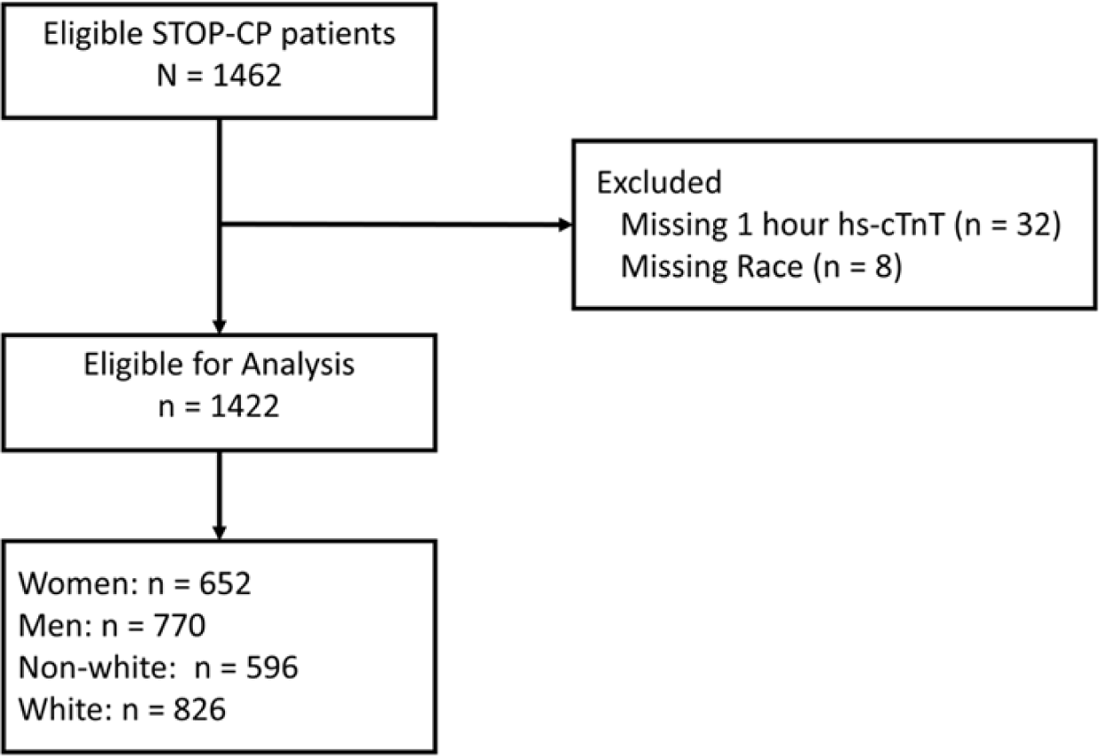
Patient flow diagram.

**Table 1.**
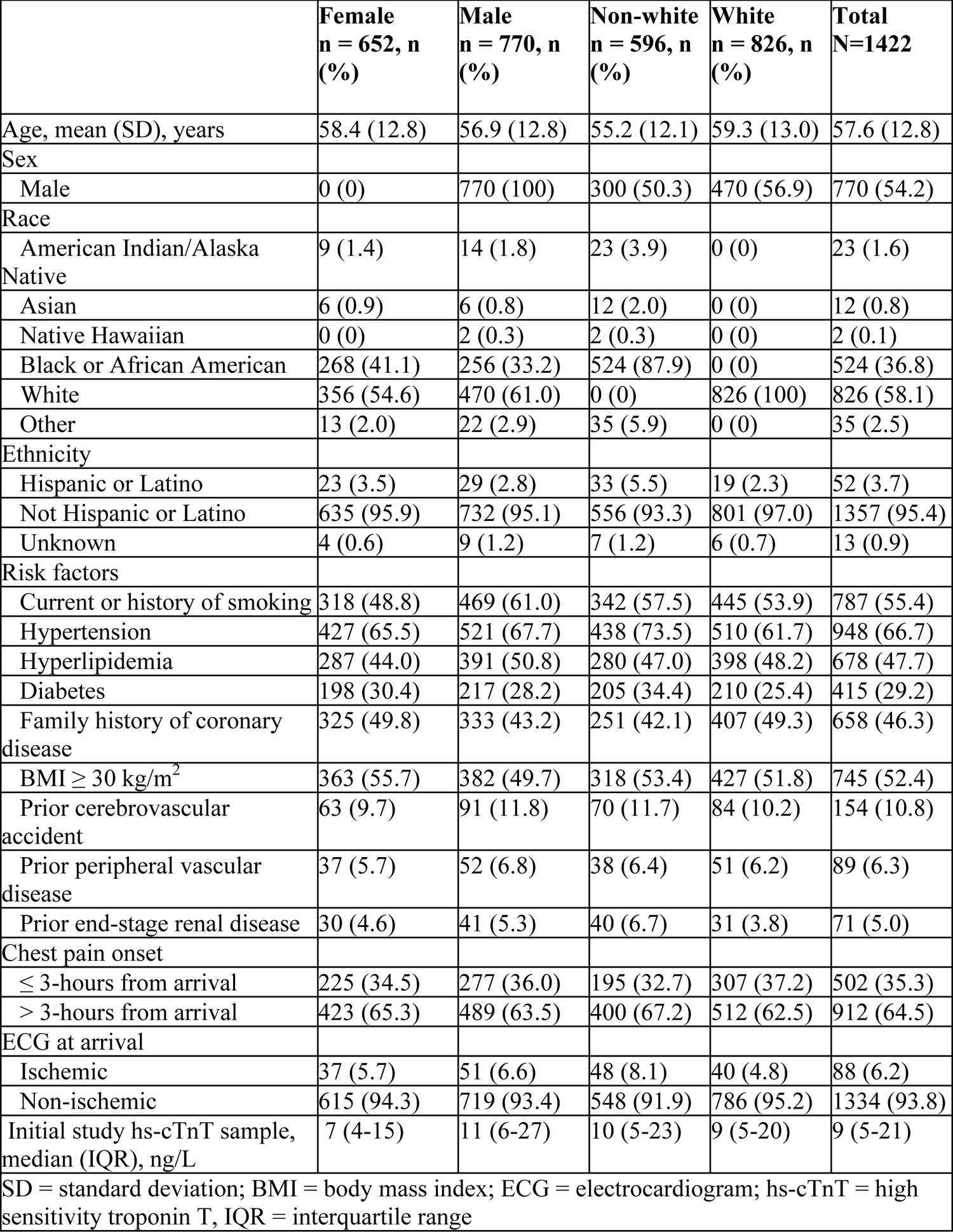
Cohort characteristics

|  | **Female**<br>**n = 652, n**<br>**(%)** | **Male**<br>**n = 770, n**<br>**(%)** | **Non-white**<br>**n = 596, n**<br>**(%)** | **White**<br>**n = 826, n**<br>**(%)** | **Total**<br>**N=1422** |
| --- | --- | --- | --- | --- | --- |
| Age, mean (SD), years | 58.4 (12.8) | 56.9 (12.8) | 55.2 (12.1) | 59.3 (13.0) | 57.6 (12.8) |
| Sex |  |  |  |  |  |
| Male | 0 (0) | 770 (100) | 300 (50.3) | 470 (56.9) | 770 (54.2) |
| Race |  |  |  |  |  |
| American Indian/Alaska Native | 9 (1.4) | 14 (1.8) | 23 (3.9) | 0 (0) | 23 (1.6) |
| Asian | 6 (0.9) | 6 (0.8) | 12 (2.0) | 0 (0) | 12 (0.8) |
| Native Hawaiian | 0 (0) | 2 (0.3) | 2 (0.3) | 0 (0) | 2 (0.1) |
| Black or African American | 268 (41.1) | 256 (33.2) | 524 (87.9) | 0 (0) | 524 (36.8) |
| White | 356 (54.6) | 470 (61.0) | 0 (0) | 826 (100) | 826 (58.1) |
| Other | 13 (2.0) | 22 (2.9) | 35 (5.9) | 0 (0) | 35 (2.5) |
| Ethnicity |  |  |  |  |  |
| Hispanic or Latino | 23 (3.5) | 29 (2.8) | 33 (5.5) | 19 (2.3) | 52 (3.7) |
| Not Hispanic or Latino | 635 (95.9) | 732 (95.1) | 556 (93.3) | 801 (97.0) | 1357 (95.4) |
| Unknown | 4 (0.6) | 9 (1.2) | 7 (1.2) | 6 (0.7) | 13 (0.9) |
| Risk factors |  |  |  |  |  |
| Current or history of smoking | 318 (48.8) | 469 (61.0) | 342 (57.5) | 445 (53.9) | 787 (55.4) |
| Hypertension | 427 (65.5) | 521 (67.7) | 438 (73.5) | 510 (61.7) | 948 (66.7) |
| Hyperlipidemia | 287 (44.0) | 391 (50.8) | 280 (47.0) | 398 (48.2) | 678 (47.7) |
| Diabetes | 198 (30.4) | 217 (28.2) | 205 (34.4) | 210 (25.4) | 415 (29.2) |
| Family history of coronary disease | 325 (49.8) | 333 (43.2) | 251 (42.1) | 407 (49.3) | 658 (46.3) |
| BMI $\geq 30$ kg/m<sup>2</sup> | 363 (55.7) | 382 (49.7) | 318 (53.4) | 427 (51.8) | 745 (52.4) |
| Prior cerebrovascular accident | 63 (9.7) | 91 (11.8) | 70 (11.7) | 84 (10.2) | 154 (10.8) |
| Prior peripheral vascular disease | 37 (5.7) | 52 (6.8) | 38 (6.4) | 51 (6.2) | 89 (6.3) |
| Prior end-stage renal disease | 30 (4.6) | 41 (5.3) | 40 (6.7) | 31 (3.8) | 71 (5.0) |
| Chest pain onset |  |  |  |  |  |
| $\leq 3$-hours from arrival | 225 (34.5) | 277 (36.0) | 195 (32.7) | 307 (37.2) | 502 (35.3) |
| $> 3$-hours from arrival | 423 (65.3) | 489 (63.5) | 400 (67.2) | 512 (62.5) | 912 (64.5) |
| ECG at arrival |  |  |  |  |  |
| Ischemic | 37 (5.7) | 51 (6.6) | 48 (8.1) | 40 (4.8) | 88 (6.2) |
| Non-ischemic | 615 (94.3) | 719 (93.4) | 548 (91.9) | 786 (95.2) | 1334 (93.8) |
| Initial study hs-cTnT sample, median (IQR), ng/L | 7 (4-15) | 11 (6-27) | 10 (5-23) | 9 (5-20) | 9 (5-21) |
| SD = standard deviation; BMI = body mass index; ECG = electrocardiogram; hs-cTnT = high sensitivity troponin T, IQR = interquartile range |  |  |  |  |  |

### ESC 0/1-Hour Algorithm by Sex

The efficacy of the hs-cTnT ESC 0/1-hour algorithm was higher in women than men, with 66.9% (436/652) of women stratified to the rule-out zone compared to 50.0% (385/770) of men (p<0.001). Among patients stratified into the rule-out zone, 30-day cardiac death or MI occurred in 1.1% (5/436) of women vs 2.1% (8/385) of men [OR 0.55, 95%CI 0.16-1.65]. MACE at 30 days did not significantly differ by sex among those in the rule-out zone, with a MACE rate of 1.8% (8/436) for women and 3.9% (15/385) for men [OR 0.46, 95%CI 0.18-1.07]. The NPV for 30-day cardiac death or MI was 98.9% (95%CI 97.3-99.6) in women and 97.9% (95%CI 95.9-99.1) in men (p=0.40). Events for men and women stratified by ESC 0/1-hour rule-out, observation, and rule-in zones are summarized in Table 2.

**Table 2.** Safety events among females and males by ESC 0/1 zone (unadjusted)

| **<u>RULE-OUT</u>** | Female n =<br>436 ,n (%) | Male n =<br>385, n (%) | Total n =<br>821, n (%) | Odds Ratio (95%<br>CI) |
| --- | --- | --- | --- | --- |
| **Index** |  |  |  |  |
| Cardiac Death | 1 (0.2) | 0 (0) | 1 (0.1) | NA |
| MI | 4 (0.9) | 4 (1) | 8 (1) | 0.88 (0.21-3.75) |
| Revascularization | 2 (0.5) | 8 (2.1) | 10 (1.2) | **0.22 (0.03-0.87)** |
| Cardiac Death or MI | 5 (1.1) | 4 (1) | 9 (1.1) | 1.1 (0.29-4.49) |
| MACE | 7 (1.6) | 9 (2.3) | 16 (1.9) | 0.68 (0.24-1.85) |
| **30-day (Index + Follow-up)** |  |  |  |  |
| Cardiac Death | 1 (0.2) | 0 (0) | 1 (0.1) | NA |
| MI | 4 (0.9) | 8 (2.1) | 12 (1.5) | 0.44 (0.12-1.4) |
| Revascularization | 3 (0.7) | 10 (2.6) | 13 (1.6) | **0.26 (0.06-0.86)** |
| Cardiac Death or MI | 5 (1.1) | 8 (2.1) | 13 (1.6) | 0.55 (0.16-1.65) |
| MACE | 8 (1.8) | 15 (3.9) | 23 (2.8) | 0.46 (0.18-1.07) |
| **<u>OBSERVATION</u>** | Female n =<br>159, n (%) | Male n =<br>252, n (%) | Total N =<br>411, n (%) | Odds Ratio (95%<br>CI) |
| **Index** |  |  |  |  |
| Cardiac Death | 0 (0) | 0 (0) | 0 (0) | NA |
| MI | 13 (8.2) | 38 (15.1) | 51 (12.4) | **0.5 (0.25-0.95)** |
| Revascularization | 4 (2.5) | 16 (6.3) | 20 (4.9) | 0.38 (0.11-1.06) |
| Cardiac Death or MI | 13 (8.2) | 38 (15.1) | 51 (12.4) | **0.5 (0.25-0.95)** |
| MACE | 13 (8.2) | 42 (16.7) | 55 (13.4) | **0.45 (0.22-0.84)** |
| **30-day (Index + Follow-up)** |  |  |  |  |
| Cardiac Death | 1 (0.6) | 1 (0.4) | 2 (0.5) | NA |
| MI | 15 (9.4) | 42 (16.7) | 57 (13.9) | **0.52 (0.27-0.95)** |
| Revascularization | 8 (5) | 21 (8.3) | 29 (7.1) | 0.58 (0.24-1.3) |
| Cardiac Death or MI | 16 (10.1) | 43 (17.1) | 59 (14.4) | **0.54 (0.29-0.98)** |
| MACE | 19 (11.9) | 50 (19.8) | 69 (16.8) | **0.55 (0.3-0.96)** |
| **<u>RULE-IN</u>** | Female n =<br>57, n (%) | Male n =<br>133, n (%) | Total N =<br>190, n (%) | Odds Ratio (95%<br>CI) |
| **Index** |  |  |  |  |
| Cardiac Death | 0 (0) | 1 (0.8) | 1 (0.5) | NA |
| MI | 35 (61.4) | 70 (52.6) | 105 (55.3) | 1.43 (0.76-2.72) |
| Revascularization | 13 (22.8) | 25 (18.8) | 38 (20) | 1.28 (0.59-2.69) |
| Cardiac Death or MI | 35 (61.4) | 70 (52.6) | 105 (55.3) | 1.43 (0.76-2.72) |
| MACE | 35 (61.4) | 70 (52.6) | 105 (55.3) | 1.43 (0.76-2.72) |
| **30-day (Index + Follow-up)** |  |  |  |  |
| Cardiac Death | 0 (0) | 6 (4.5) | 6 (3.2) | NA |
| MI | 36 (63.2) | 73 (54.9) | 109 (57.4) | 1.41 (0.75-2.69) |
| Revascularization | 15 (26.3) | 28 (21.1) | 43 (22.6) | 1.34 (0.64-2.73) |
| Cardiac Death or MI | 36 (63.2) | 75 (56.4) | 111 (58.4) | 1.33 (0.7-2.54) |
| MACE | 36 (63.2) | 75 (56.4) | 111 (58.4) | 1.33 (0.7-2.54) |

The ESC 0/1-hour algorithm classified 8.7% (57/652) of women into the rule-in zone vs. 17.3% of men (133/770) (p<0.001). Among rule-in patients, the rate of index cardiac death or MI was similar between women and men [61.4% (35/57) vs. 52.6% (70/133); OR 1.43, 95%CI 0.76-2.72]. The PPV for 30-day cardiac death or MI was 63.2% (95%CI 49.3-75.6) for women and 56.4% (95%CI 47.5-65.0) for men (p=0.42). However, the +LR for 30-day cardiac death or MI was higher for women than men (17.9, 95%CI 11.3-28.5 vs. 6.6, 95%CI 5.0-8.8; p< 0.001). The diagnostic performance of the algorithm by sex and race is summarized in Table 3 and Figure 2.

**Table 3.** Test characteristics of ESC 0/1

|  |  | 30-day Cardiac Death or MI |  |  |  |
| --- | --- | --- | --- | --- | --- |
|  |  | Females | Males | Non-whites | Whites |
| **Rule-Out** | Sensitivity (95%CI) | 91.2 (80.7-97.1) | 93.7 (87.9-97.2) | 94.0 (85.4-98.3) | 92.2 (85.8-96.4) |
|  | NPV (95%CI) | 98.9 (97.3-99.6) | 97.9 (95.9-99.1) | 98.8 (96.9-99.7) | 98.2 (96.5-99.2) |
|  | -LR (95%CI) | 0.12 (0.05-0.28) | 0.11 (0.06-0.21) | 0.10 (0.04-0.25) | 0.12 (0.06-0.22) |
| **Rule-In** | Specificity (95%CI) | 96.5 (94.7-97.8) | 91.0 (88.5-93.1) | 91.1 (88.4-93.4) | 95.5 (93.7-96.9) |
|  | PPV (95%CI) | 63.2 (49.3-75.6) | 56.4 (47.5-65.0) | 49.5 (38.9-60.0) | 67.0 (56.7-76.2) |
|  | +LR (95%CI) | 17.9 (11.3-28.5) | 6.6 (5.0-8.8) | 7.7 (5.6-10.6) | 12.4 (8.5-18.1) |
|  |  | 30-day MACE |  |  |  |
|  |  | Females | Males | Non-whites | Whites |
| **Rule-Out** | Sensitivity (95%CI) | 87.3 (76.5-94.4) | 89.3 (82.9-93.9) | 90.1 (80.7-95.9) | 87.9 (81.1-92.9) |
|  | NPV (95%CI) | 98.2 (96.4-99.2) | 96.1 (93.7-97.8) | 97.9 (95.7-99.1) | 96.7 (94.8-98.1) |
|  | -LR (95%CI) | 0.18 (0.09-0.34) | 0.18 (0.11-0.30) | 0.16 (0.08-0.32) | 0.18 (0.11-0.28) |
| **Rule-In** | Specificity (95%CI) | 96.4 (94.6-97.8) | 90.8 (88.3-92.9) | 91.0 (88.3-93.3) | 95.4 (93.6-96.8) |
|  | PPV (95%CI) | 63.2 (49.3-75.6) | 56.4 (47.5-65.0) | 49.5 (38.9-60.0) | 67.0 (56.7-76.2) |
|  | +LR (95%CI) | 16.0 (10.0-25.7) | 5.8 (4.4-7.8) | 7.2 (5.2-10.0) | 10.7 (7.3-15.6) |

**Figure 2A-B.**
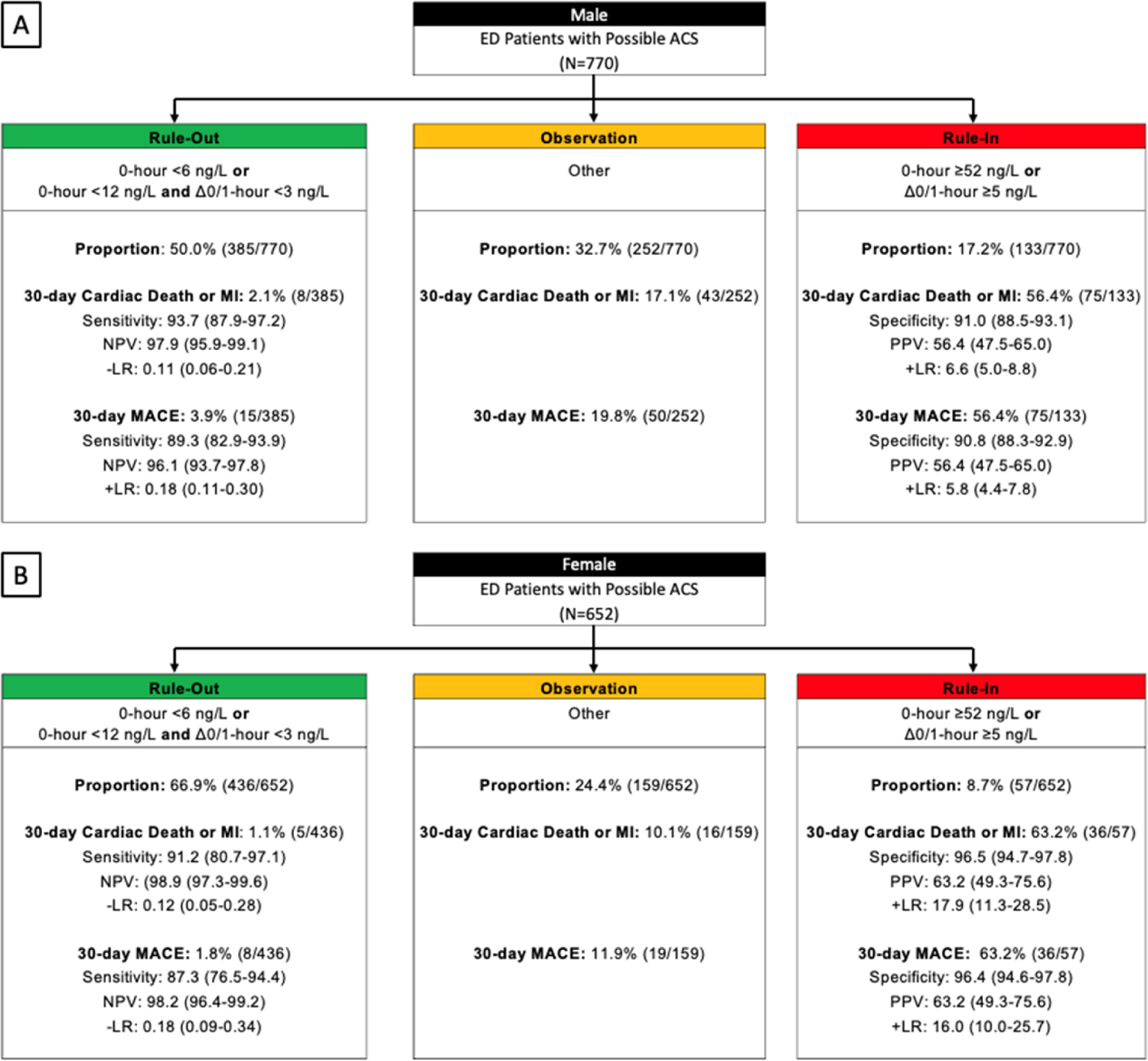
The ESC 0/1-hour algorithm among male and female patients for 30-day cardiac death or MI, and 30-day MACE. A – Male, B – Female ACS – acute coronary syndrome, CAD – coronary artery disease, ED – emergency department; MI – myocardial infarction; MACE – major adverse cardiovascular event; NPV – negative predictive value, PPV – positive predictive value, LR – likelihood ratio

The interaction between the ESC 0/1-h algorithm and sex was significant for 30-day cardiac death or MI (p=0.04) and 30-day MACE (p=0.021). The adjusted odds of 30-day cardiac death or MI and 30-day MACE were higher for men in the rule-out and observation zones based on point estimates, but higher for women in the rule-in zone. Figure 3 and Supplemental Table 1 show the aORs for index and 30-day cardiac death or MI and MACE.

**Figure 3A-B.**
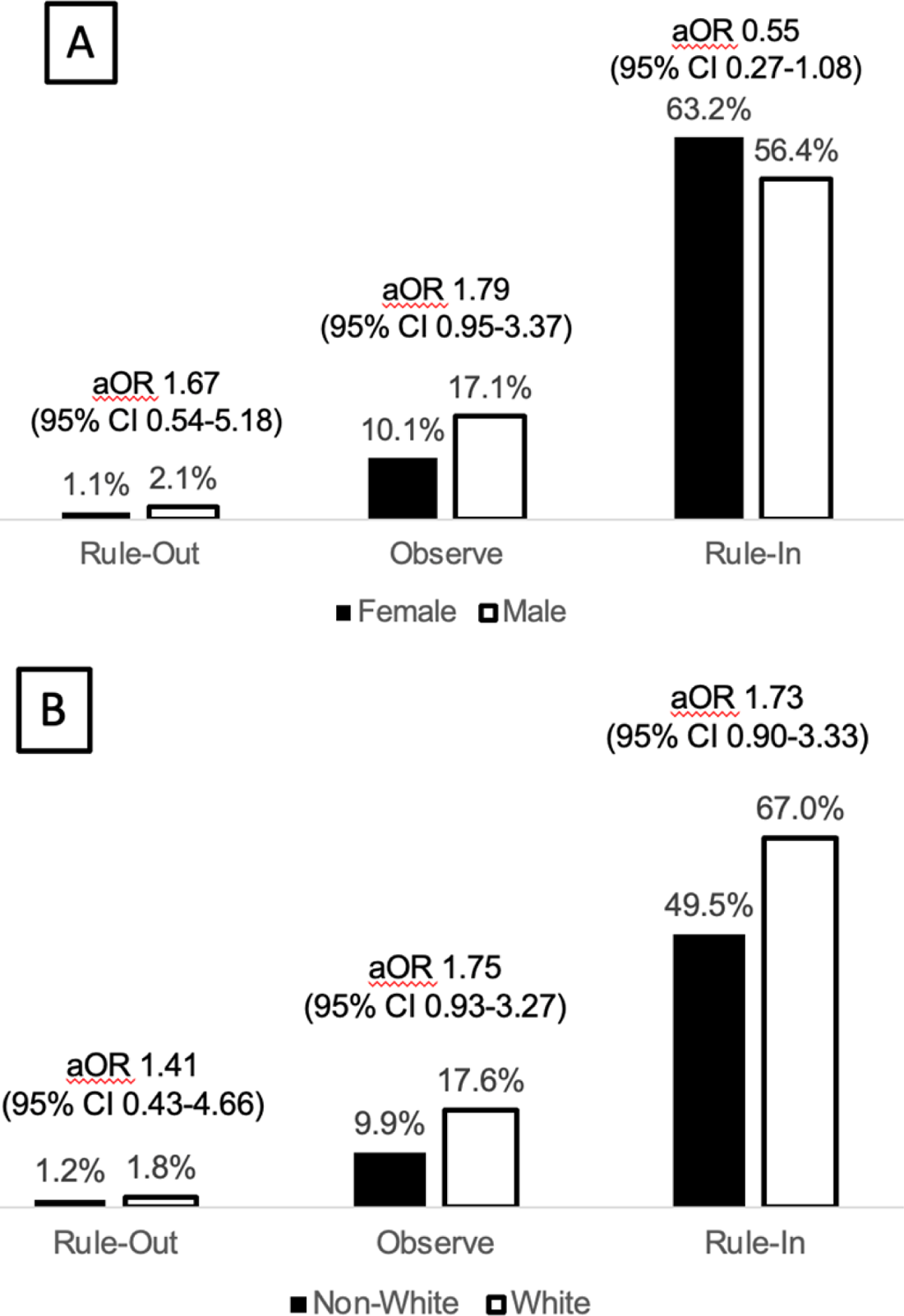
Proportion of patients with 30-day cardiac death or MI by ESC0/1 category. A – Sex, B – Race ACS – acute coronary syndrome, CAD – coronary artery disease, ED – emergency department; MI – myocardial infarction; NPV – negative predictive value, PPV – positive predictive value, LR – likelihood ratio, aOR – adjusted odds ratio.

### ESC 0/1-Hour Algorithm by Race

The efficacy of the ESC 0/1-hour algorithm was similar between non-white and white patients [55.5% (331/596) vs. 59.3% (490/826); p = 0.16]. Among patients who were ruled out by ESC 0/1-hour algorithm, cardiac death or MI at 30-days occurred in 1.2% (4/331) of non-white patients compared to 1.8% (9/490) of white patients [OR 0.65, 95%CI 0.18-2.03]. MACE at 30-days was also similar by race among patients classified into the rule-out zone, with a MACE rate of 2.1% (7/331) for non-white patients compared to 3.3% (16/490) for white patients [OR 0.64, 95%CI 0.24-1.52]. The NPV for 30-day cardiac death or MI was 98.8% (95%CI 96.9-99.7) for non-white patients and 98.2% (96.5-99.2) for white patients (p=0.58). Event rates for non-white and white patients stratified by ESC 0/1-hour algorithm rule-out, observation, and rule-in zones are summarized in Table 4 and Figure 4.

**Table 4.**
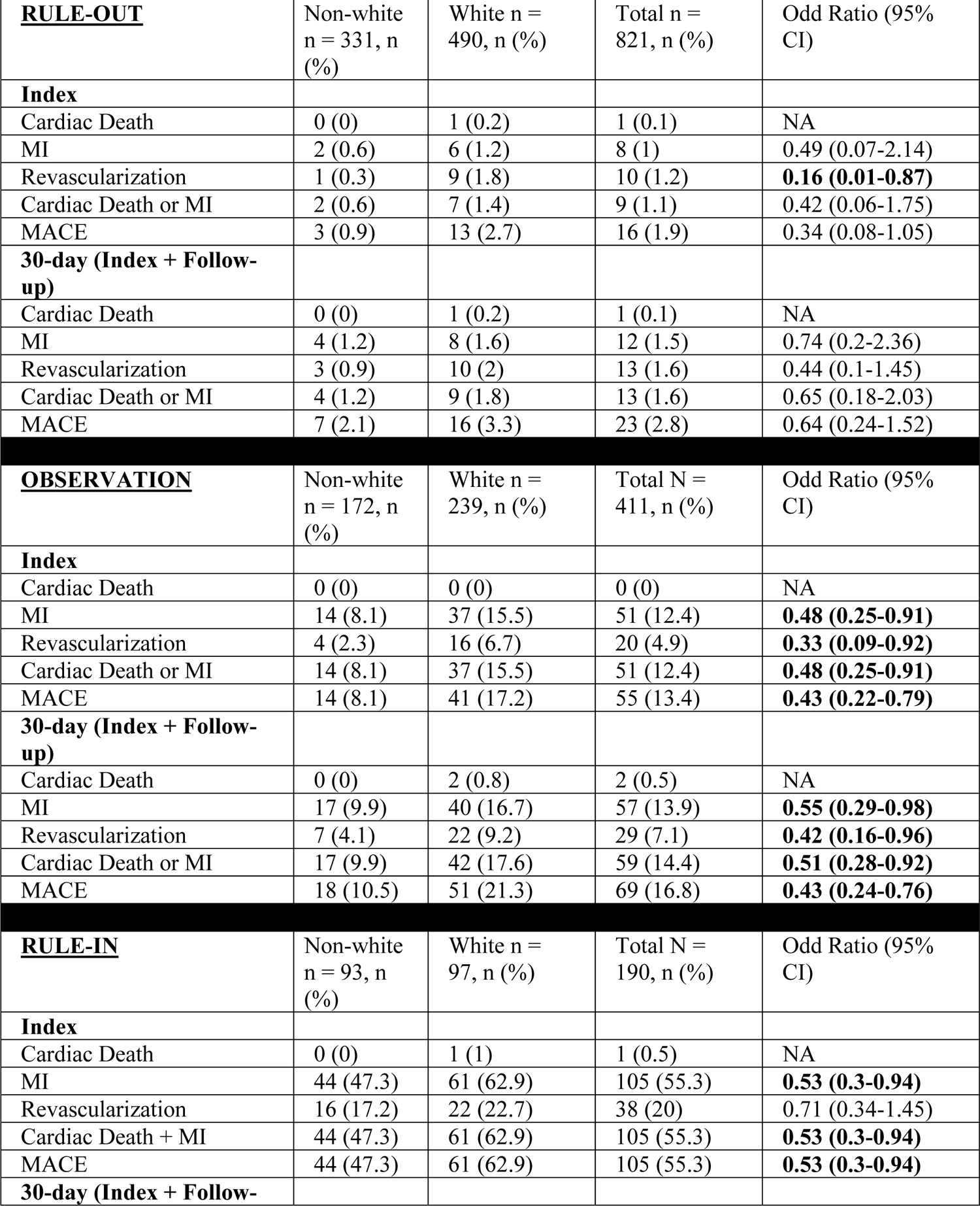
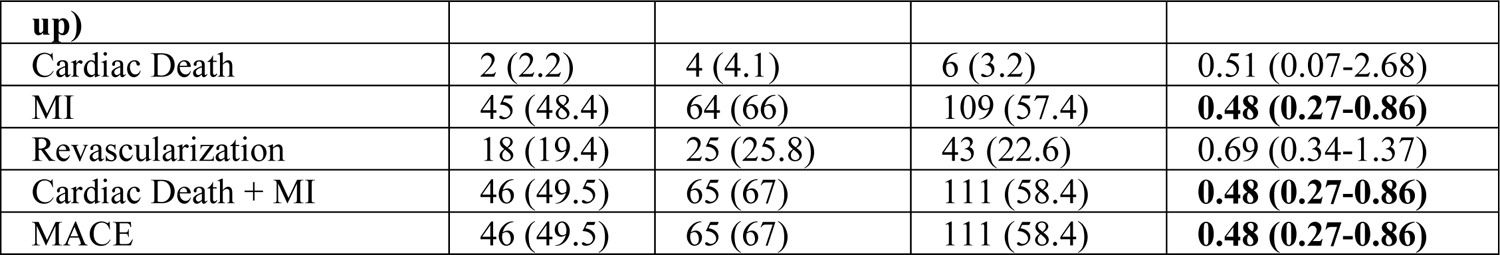
Safety events among non-white and white patients by ESC 0/1 zone (unadjusted)

| **<u>RULE-OUT</u>** | Non-white<br>n = 331, n<br>(%) | White n =<br>490, n (%) | Total n =<br>821, n (%) | Odd Ratio (95%<br>CI) |
| --- | --- | --- | --- | --- |
| **Index** |  |  |  |  |
| Cardiac Death | 0 (0) | 1 (0.2) | 1 (0.1) | NA |
| MI | 2 (0.6) | 6 (1.2) | 8 (1) | 0.49 (0.07-2.14) |
| Revascularization | 1 (0.3) | 9 (1.8) | 10 (1.2) | **0.16 (0.01-0.87)** |
| Cardiac Death or MI | 2 (0.6) | 7 (1.4) | 9 (1.1) | 0.42 (0.06-1.75) |
| MACE | 3 (0.9) | 13 (2.7) | 16 (1.9) | 0.34 (0.08-1.05) |
| **30-day (Index + Follow-up)** |  |  |  |  |
| Cardiac Death | 0 (0) | 1 (0.2) | 1 (0.1) | NA |
| MI | 4 (1.2) | 8 (1.6) | 12 (1.5) | 0.74 (0.2-2.36) |
| Revascularization | 3 (0.9) | 10 (2) | 13 (1.6) | 0.44 (0.1-1.45) |
| Cardiac Death or MI | 4 (1.2) | 9 (1.8) | 13 (1.6) | 0.65 (0.18-2.03) |
| MACE | 7 (2.1) | 16 (3.3) | 23 (2.8) | 0.64 (0.24-1.52) |
| **<u>OBSERVATION</u>** | Non-white<br>n = 172, n<br>(%) | White n =<br>239, n (%) | Total N =<br>411, n (%) | Odd Ratio (95%<br>CI) |
| **Index** |  |  |  |  |
| Cardiac Death | 0 (0) | 0 (0) | 0 (0) | NA |
| MI | 14 (8.1) | 37 (15.5) | 51 (12.4) | **0.48 (0.25-0.91)** |
| Revascularization | 4 (2.3) | 16 (6.7) | 20 (4.9) | **0.33 (0.09-0.92)** |
| Cardiac Death or MI | 14 (8.1) | 37 (15.5) | 51 (12.4) | **0.48 (0.25-0.91)** |
| MACE | 14 (8.1) | 41 (17.2) | 55 (13.4) | **0.43 (0.22-0.79)** |
| **30-day (Index + Follow-up)** |  |  |  |  |
| Cardiac Death | 0 (0) | 2 (0.8) | 2 (0.5) | NA |
| MI | 17 (9.9) | 40 (16.7) | 57 (13.9) | **0.55 (0.29-0.98)** |
| Revascularization | 7 (4.1) | 22 (9.2) | 29 (7.1) | **0.42 (0.16-0.96)** |
| Cardiac Death or MI | 17 (9.9) | 42 (17.6) | 59 (14.4) | **0.51 (0.28-0.92)** |
| MACE | 18 (10.5) | 51 (21.3) | 69 (16.8) | **0.43 (0.24-0.76)** |
| **<u>RULE-IN</u>** | Non-white<br>n = 93, n<br>(%) | White n =<br>97, n (%) | Total N =<br>190, n (%) | Odd Ratio (95%<br>CI) |
| **Index** |  |  |  |  |
| Cardiac Death | 0 (0) | 1 (1) | 1 (0.5) | NA |
| MI | 44 (47.3) | 61 (62.9) | 105 (55.3) | **0.53 (0.3-0.94)** |
| Revascularization | 16 (17.2) | 22 (22.7) | 38 (20) | 0.71 (0.34-1.45) |
| Cardiac Death + MI | 44 (47.3) | 61 (62.9) | 105 (55.3) | **0.53 (0.3-0.94)** |
| MACE | 44 (47.3) | 61 (62.9) | 105 (55.3) | **0.53 (0.3-0.94)** |
| **30-day (Index + Follow-** |  |  |  |  |
| **up)** |  |  |  |  |
| Cardiac Death | 2 (2.2) | 4 (4.1) | 6 (3.2) | 0.51 (0.07-2.68) |
| MI | 45 (48.4) | 64 (66) | 109 (57.4) | **0.48 (0.27-0.86)** |
| Revascularization | 18 (19.4) | 25 (25.8) | 43 (22.6) | 0.69 (0.34-1.37) |
| Cardiac Death + MI | 46 (49.5) | 65 (67) | 111 (58.4) | **0.48 (0.27-0.86)** |
| MACE | 46 (49.5) | 65 (67) | 111 (58.4) | **0.48 (0.27-0.86)** |

**Figure 4A-B.**
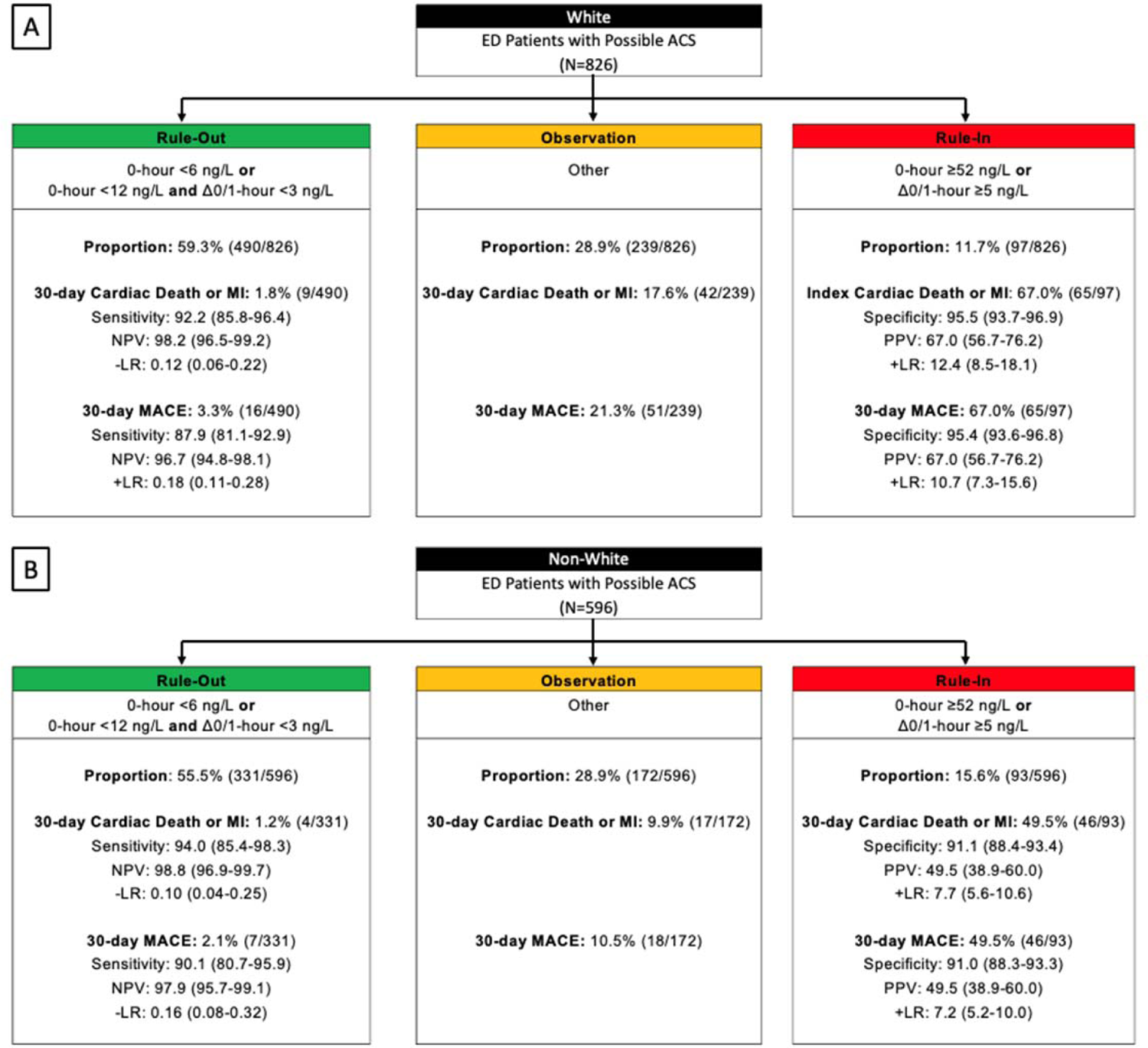
The ESC 0/1-hour algorithm among white and non-white patients for 30-day cardiac death or MI and 30-day MACE. A – White, B – Non-White ACS – acute coronary syndrome, CAD – coronary artery disease, ED – emergency department; MI – myocardial infarction; MACE – major adverse cardiovascular event; NPV – negative predictive value, PPV – positive predictive value, LR – likelihood ratio

The ESC 0/1-hour algorithm classified 15.6% (93/596) of non-white patients into the rule-in zone vs. 11.7% (97/826) of white patients (p=0.04). Among rule-in patients, the rate of index cardiac death or MI was lower in non-white patients than white patients [47.3% (44/93) vs. 62.9% (61/97); OR 0.53, 95%CI 0.30-0.94]. At 30 days, 49.5% (46/93) of non-white patients experienced cardiac death or MI compared to 67.0% (65/97) of white patients (OR 0.48, 95%CI 0.27-0.86). The PPV for 30-day cardiac death or MI was 49.5% (95%CI 38.9-60.0) for non-white patients and 67.0% (95%CI 56.7-76.2) for white patients (p=0.21). The test characteristics for the rule-in zone are presented in Table 3.

The overall interaction of the ESC 0/1-hour algorithm with race was not significant for outcomes of 30-day cardiac death or MI (p=0.951) or 30-day MACE (p=0.861). Adjusted odds ratio point estimates remained greater than one for white patients compared to non-white patients in each ESC 0/1-hour algorithm zone. Figure 3 and Supplemental Table 1 show the aORs for index and 30-day cardiac death or MI and MACE.

## Discussion

In this multicenter, prospective study, the hs-cTnT ESC 0/1-hour algorithm failed to achieve an acceptable^15^ NPV ≥99% for 30-day cardiac death or MI among any sex or race subgroup. Although efficacy was higher in women, there was no difference in safety among men vs. women. While fewer women were placed in the rule-in zone, they were more likely to experience 30-day cardiac death or MI or MACE than men. Efficacy and safety were similar among white and non-white patients. More non-white patients were classified to the rule-in zone, but they had lower odds of 30-day cardiac death or MI than white patients in the rule-in zone.

Use of the hs-cTnT ESC 0/1-hour algorithm for evaluation of ED patients with chest pain is a class I recommendation by the European Society of Cardiology guidelines.^4^ This recommendation is based on international studies of the algorithm, which have demonstrated a NPV ≥99% for index-MI,^32, 33^ 30-day cardiac death or MI,^34^ or 30-day MACE.^6, 7^ However, in some cohorts the hs-cTnT ESC 0/1-hour algorithm has failed to achieve the ≥ 99% NPV for adverse cardiac events, which is the threshold most physicians consider acceptable.^30, 35–39^ This includes the primary analysis of the U.S.-based STOP-CP trial.^20^ Data from this subgroup analysis demonstrates adds to that primary analysis by demonstrating that the algorithm failed to achieve a NPV ≥99% for 30-day cardiac death or MI or 30-day MACE among any sex or race subgroup. This suggests the hs-cTnT ESC 0/1-hour algorithm alone may not be able to adequately identify US ED patients with chest pain who are safe for discharge.

Our results demonstrated that the hs-cTnT ESC 0/1-hour algorithm placed more women into the rule-out zone compared to men. This is consistent with a prior subgroup analysis of the hs-cTnT 0/1-hour algorithm in European and U.S. cohorts.^6, 7, 14, 39^ Sex-based differences in efficacy may be explained by sex-specific differences in normal troponin ranges and thresholds for MI diagnosis. Previous evidence suggests that normal troponin concentrations are lower among women compared to men, possibly due to differences in left ventricular mass.^16, 40, 41^ The Fourth Universal Definition of Myocardial Infarction (UDMI) recommends using sex-specific troponin cutoffs. When used, sex-specific hs-cTnT 99^th^ percentile URL cutoffs increase the diagnosis of MI among women.^16, 27^ Kimenai et. al, found that at given levels of elevated hs-cTnT, women had higher risk of cardiovascular events than men, which may explain why we found a higher point estimate odds of 30-day cardiac death or MI for women in the rule-in group compared to men.^42^ We did not observe increased rates of 30-day cardiac death or MI or MACE for women relative to men in the rule-out zone, but the study may have been underpowered to detect this difference. The higher proportion of 30-day cardiac death or MI among women in the rule-in zone compared to men suggests sex-specific rule-in hs-cTnT thresholds may be needed for chest pain diagnostic pathways and should be a point of future study.

We found no significant interaction between race and ESC 0/1-hour algorithm for 30-day cardiac death or MI, with white patients in each ESC 0/1-hour algorithm zone having higher odds of 30-day cardiac death or MI compared to non-white patients. However, despite a significantly higher proportion of non-white patients placed in the rule-in zone compared to white patients, non-white patients had significantly lower odds of 30-day cardiac death or MI compared to white patients in the rule-in zone. This suggests that use of hs-cTnT ESC 0/1-hour algorithm may lead to disproportionate over-testing among non-white patients. The underlying mechanisms for this are unclear and warrant further study but may be related to unmeasured social determinants of health,. In addition, future studies are needed to determine if integrating individualized hs-cTn cutoffs based on race, social determinants, or other variables into a 0/1-hour algorithm reduces over-classification of patients into the rule-in zone.

### Limitations

Although this study was conducted at eight U.S. EDs, these were mostly academic sites, which limits generalizability to other care settings. Informed consent was required to participate in STOP-CP, resulting in possible selection bias. Sex and race were determined by patient self-reporting and medical record review which may have led to misclassification bias. Ethnicity was not included in this analysis. The 30-day cardiac death or MI and MACE rates in STOP-CP are higher than in previous U.S. cohorts, and this increased prevalence may impact NPV and PPV.^14, 30^ As required by the Food and Drug Administration, the lowest reportable value for the hs-cTnT assay was 6 ng/L, which is different than the ESC 0/1-hour algorithm rule-out threshold hs-cTnT value of 5 ng/L used outside of the U.S. Our prior analyses suggest that this change has a minimal impact on the performance of the hs-cTnT ESC 0/1-hour algorithm.^43, 44^ However, it may affect the generalizability of these results outside of the U.S. This study used only the Roche hs-cTnT assay. Therefore, these conclusions cannot be applied to ESC 0/1-hour hs-cTnI algorithm derivations. Finally, this study was observational and as such, the ESC 0/1-hour hs-cTnT algorithm was not used to guide patient care.

## Conclusions

In this multisite, prospective U.S. cohort study, the hs-cTnT ESC 0/1-hour algorithm did not achieve a NPV ≥ 99% in any sex or race subgroup. Efficacy was significantly higher for women compared to men. More non-white patients were classified into the rule-in zone, though they had significantly lower adjusted odds of 30-day cardiac death or MI compared to white patients in the rule-in zone. Future studies should assess the integration of individualized hs-cTnT thresholds to optimize risk stratification of ED patients with chest pain.

## Data Availability

Data

## Acknowledgements

We appreciate Mate Huis in’t Veld MD; Michael Massoomi MD; and James McCord MD for adjudicating outcomes in the STOP-CP study. We appreciate Brennan E. Paradee MS for her contribution to the analysis.

## SUPPLEMENTAL MATERIALS

**eTable 1.** Adjusted odds ratios (aOR) for safety events among ESC 0/1-hour hs-cTnT algorithm patients who were male vs. female and white vs. non-white.

| **Rule-Out** | **aOR (95%CI)<sup>a</sup> Sex** | **aOR (95%CI)<sup>a</sup> Race** |
| --- | --- | --- |
| Index Cardiac Death or MI | 0.82 (0.22-3.10) | 2.17 (0.45-10.5) |
| Index MACE | 1.29 (0.47-3.52) | 2.71 (0.76-9.67) |
| 30-day Cardiac Death or MI | 1.67 (0.54-5.18) | 1.41 (0.43-4.66) |
| 30-day MACE | 1.85 (0.77-4.47) | 1.47 (0.59-3.64) |
| **Observation** |  |  |
| Index Cardiac Death or MI | 1.86 (0.94-3.68) | 1.81 (0.92-3.57) |
| Index MACE | 2.08 (1.06-4.09) | 2.05 (1.05-4.00) |
| 30-day Cardiac Death or MI | 1.79 (0.95-3.37) | 1.75 (0.93-3.27) |
| 30-day MACE | 1.72 (0.95-3.12) | 2.15 (1.18-3.94) |
| **Rule-In** |  |  |
| Index Cardiac Death or MI | 0.51 (0.25-1.05) | 1.62 (0.85-3.10) |
| Index MACE | 0.49 (0.24-1.01) | 1.54 (0.81-2.95) |
| 30-day Cardiac Death or MI | 0.55 (0.27-1.12) | 1.73 (0.90-3.33) |
| 30-day MACE | 0.52 (0.26-1.08) | 1.73 (0.90-3.33) |

MI – myocardial infarction, MACE – major adverse cardiovascular event, CAD – coronary artery disease
<sup>a</sup> Adjustment was made for age, sex, race, hypertension, diabetes, hyperlipidemia, obesity, current smoking, prior stroke, peripheral vascular disease, and end stage renal disease as well as the interaction between ESC 0/1-hour classification and known CAD.

